# Clinical profile and immediate outcome of Multisystem Inflammatory Syndrome in Children (MIS-C) associated with Covid-19: a multicentric study

**DOI:** 10.1101/2021.06.21.21259259

**Authors:** Geetanjali Sethy, Bibhudatta Mishra, Mukesh Kumar Jain, Sibabratta Patnaik, Reshmi Mishra, Jyoti Ranjan Behera, Bandya Sahoo

## Abstract

**INTRODUCTION:** Following an asymptomatic or mildly symptomatic Corona virus disease (COVID 19), otherwise healthy children, may develop serious manifestations in form of cardiac, neurological, respiratory, gastrointestinal and dermatologic dysfunction. Many such cases were being observed in Odisha, an eastern state of India and reported from different health care facilities. We related these unexplained serious manifestations to Multisystem Inflammatory Syndrome associated with COVID 19 (MIS-C) and planned this study.

**METHODS:** This retrospective observational study was carried out in three tertiary care centres: Kalinga Institute of Medical Sciences, Bhubaneswar, MKCG Medical college Berhampur and Jagannath Hospital, Bhubaneswar between July to September of year 2020. Study population include all children from 1 month to 15 years admitted to hospital with MIS-C according to WHO Diagnostic Criteria. All the data were analyzed by SPSS.

**RESULTS:** A total of 21 children were included in our study. Maximum number of cases were male (76.2%), predominate age group was 6-10 yrs (47.6%). Predominate symptoms /signs in our observation were fever, pain abdomen, seizure and hypotension. Most of these cases were positive for SARS CoV antibody (80.95%). Response to immunotherapy was dramatic. Mortality (9%) of our study is higher to 1.8–3% from western literature. None of our patient had coronary abnormality while 2 had mild cardiac dysfunction at discharge comparable to other studies.

**CONCLUSION:** MIS-C following exposure to COVID 19 infection in children is a clinical syndrome which needs early suspicion and appropriate intervention to prevent mortality.

## INTRODUCTION

Severe acute respiratory syndrome coronavirus 2 (SARS-CoV-2) is a novel coronavirus which continues to spread and remains a threat to human life across the globe. The pediatric population infected with the virus are usually asymptomatic or exhibit mild symptoms. Multisystem inflammatory syndrome in children (MIS-C) is a new dangerous childhood disease that occurs weeks after a mild or asymptomatic SARS-CoV-2 infection. It was first reported in April, 2020. This occurs 3 to 4 weeks after an asymptomatic infection. Otherwise healthy children may manifest some combinations of cardiac dysfunction, gastrointestinal distress, fever, fatigue, or rash. This is postulated to be related to systemic inflammatory response due to COVID-19. This rare syndrome shares common features with other paediatric inflammatory conditions including: Kawasaki disease, staphylococcal and streptococcal toxic shock syndromes, bacterial sepsis and macrophage activation syndromes. It can also present with unusual abdominal symptoms, neurological symptoms (AES) with excessive inflammatory markers.

Many such cases are being observed in our state and reported from different health care facilities, we related these unexplained serious manifestations in some of the children to MIS-C. So we planned to conduct a retrospective study on these observations. In this multicentic record based study, we analyzed the data to have a clearer image of disease spectrum for early recognition and management.

## METHODS

The first case of MIS-C in Odisha was detected in first week of August 2020, corresponding to the surge of COVID 19 cases in late May and June 2020 onwards. This study was conducted after taking permission from Institutional Ethics Committee. Data were collected from medical records of three tertiary care centres of Odisha, an eastern Indian State. All the cases adhering to the WHO definition of MIS-C were included in the study. The data collected were entered in Microsoft Excel spreadsheet and imported to SPSS software. Continuous variables were expressed as means, standard deviations and categorical variables were expressed as percentages.

## RESULTS

We have seen 21 cases of MIS-C from first week of August 2020 to second week of September 2020 (Table 1). Out of them 16 (76.2%) were boys while 5 (23.8%) were girls. We found 3 (14.3%) cases between 1 to 5 years’ age group while 10 (47.6%) cases were between 6-10 yrs, 6 (28.6%) cases between 11-15 years and 2 (9.5%) cases were above 15 years. The mean age of patients was 9.09 years. Fever was found in all (100%) cases, while abdominal symptoms were found in 14 (66.7%) cases (Table 2). Loose motion, vomiting and abdominal pain were the presenting complaints in 4 (19%), 10 (47.6%) and 11 (52.4%) cases respectively. Respiratory distress at admission was found in 9 cases (42.8%), while hypotension at admission was found in 10 (47.6%) cases. Ten (47.6%) children had some form of neurological involvement like seizures, agitation or altered sensorium. Seizure was one of the manifestations in 5 (23.8%) cases. Skin rash was found in 9 (42.9%) cases while conjunctival congestion was found in 8 (38.1%). Mean total leucocyte count (TLC) was 13,800/cmm, neutrophil 72%, lymphocyte count 22% and mean platelet count was 2.40 lakh/cmm. The inflammatory markers like C-reactive protein (CRP) mean value was 52.2 mg/dL, procalcitonin level 14.9 ng/mL, while mean ESR was 76.4 mm in first hour. Mean value of ferritin was 530 ng/mL and that of LDH was 802 U/L, while mean D-dimer value was 3.832 mcg/ml. Laboratory evidence of SARS-COV-2 was found in 19 patients; 2 were RTPCR positive, 17 were IgG antibody positive while 2 had both RTPCR and antibody positive. In 2 children RTPCR and antibody were negative but their parents were RTPCR positive 4 weeks before. Out of 21 patients, 9 (42.9%) received IVIG while 20 (95.2%) received steroids, only steroid was used in 13 children (Table 3). Out of the 2 children who left against medical advice, one child did not receive any treatment. Invasive ventilation was required for 2 (0.95%) patients while non-invasive ventilation (NIV) was used for 2(0.95%) children. Inotropic support was required in 10 (47.6%) patients. Echocardiography was done in 17 cases; myocardial dysfunction was found in 8 (38.1%), while none had coronary dilatation. Out of 21 patients, 17 recovered while 2 left against medical advice and 2 died. Out of the 2 children who died, one had comorbidity like nephrotic syndrome and the other one died of respiratory complications. None of our patient had coronary abnormality while 2 had mild cardiac dysfunction at discharge.

**1. Table I.**
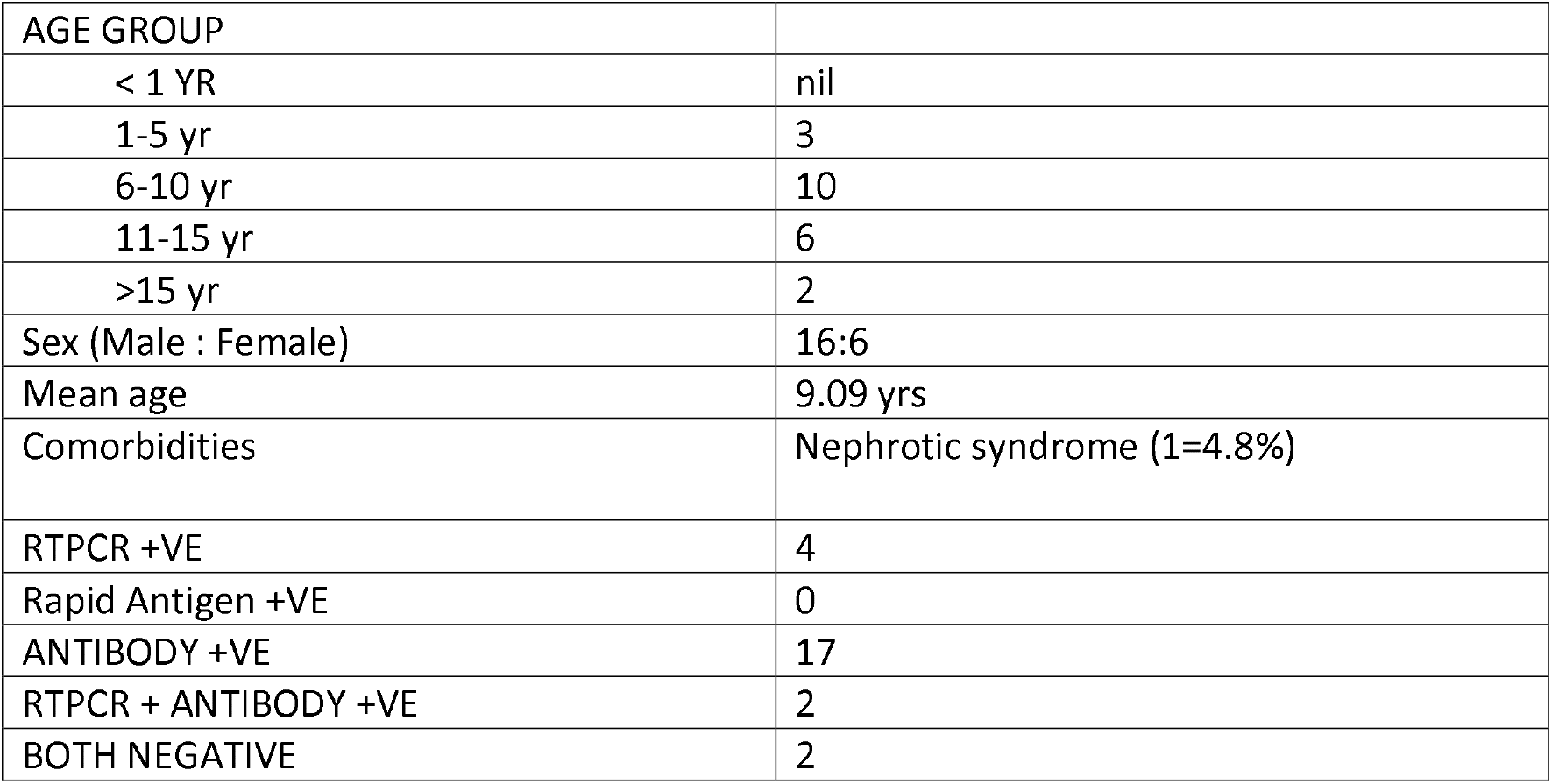
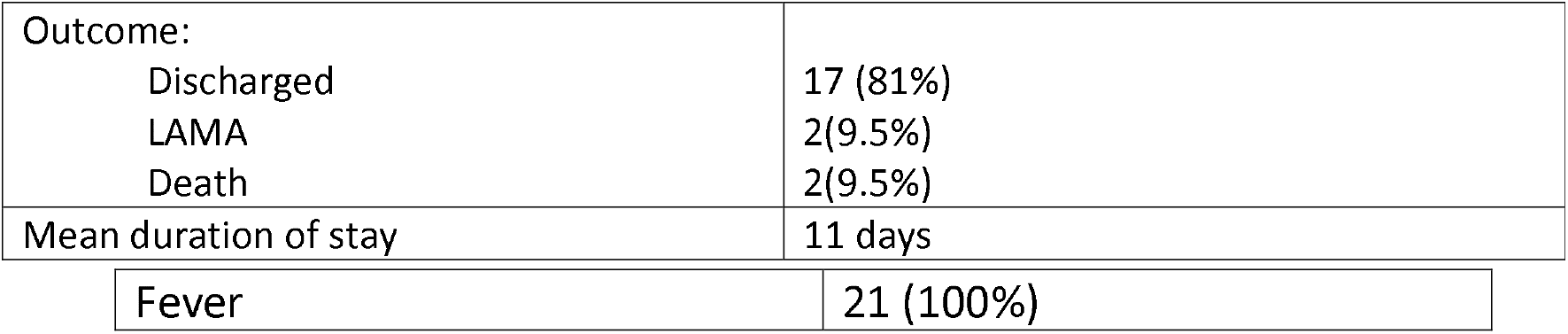
Demographic Characteristics.

|  |  |
| --- | --- |
| AGE GROUP |  |
| < 1 YR | nil |
| 1-5 yr | 3 |
| 6-10 yr | 10 |
| 11-15 yr | 6 |
| >15 yr | 2 |
| Sex (Male : Female) | 16:6 |
| Mean age | 9.09 yrs |
| Comorbidities | Nephrotic syndrome (1=4.8%) |
| RTPCR +VE | 4 |
| Rapid Antigen +VE | 0 |
| ANTIBODY +VE | 17 |
| RTPCR + ANTIBODY +VE | 2 |
| BOTH NEGATIVE | 2 |
| Outcome: |  |
| Discharged | 17 (81%) |
| LAMA | 2(9.5%) |
| Death | 2(9.5%) |
| Mean duration of stay | 11 days |
| Fever | 21 (100%) |

**TABLE: II.** Clinical characteristics

|  |  |
| --- | --- |
| Abdominal symptoms | 14 (66.7%) |
| Loose motion | 4 (19%) |
| Vomiting | 10 (47.6%) |
| Pain abdomen | 11 (52.4%) |
| Hypotension | 10 (47.6%) |
| Seizures | 5 (23.8%) |
| Respiratory Distress | 9 (42.9%) |
| Congestive heart failure | 1 (4.8%) |
| CNS involvement | 11 (47.6%) |
| Conjunctival congestion | 8 (38.1%) |
| Skin Rash | 8 (38.1%) |

**TABLE: III.** Therapeutic Options Utilised:

| Therapeutic option | Number (%) |
| --- | --- |
| IVIG | 9 (42.9%) |
| Steroid | 15 (71.4%) |
| Only steroid | 7 (33.3%) |
| Only IVIG | 1 (4.8%) |
| NIV | 2 (9.5%) |
| INVASIVE VENTILATION | 2 (9.5%) |
| LMWH | 10 (47.6%) |
| INOTROPE | 10 (47.6%) |

## DISCUSSION

Among the 21 cases, the mean age in our study population was 9.09yrs which is comparable to the mean age of 9.5yrs in a study done by Sadiq M et al in Pakistan.^1^Three cases were below 5yrs of age and 2 cases were above 15yrs, while majority (76.2%) were in between 5-15yrs age group, which points towards the vulnerable age group. Though there is a clinical resemblance towards atypical Kawasaki disease, unlike Kawasaki disease, these cases have occurred in older children and adolescents. Many studies from USA and Europe have similar age distributions like our study group.^2-5^Selva and colleagues found marked differences between the antibody responses in children and adults against corona virus. These varying responses were associated with different Fcγ receptor binding properties.^6^ Differences in antibody response might contribute to such specific age distribution in our study population.

All children in our study with features of this new inflammatory syndrome fulfilled the WHO criteria for MIS-C for the clinical, laboratory and echocardiographical features.^7^Skin rash was found in 9 (42.8%) cases while conjunctival congestion was found in 8 (38.1%). 48% of our children presented with shock and required volume resuscitation and inotropic support. Kawasaki disease-like features of the syndrome seem to be predominant in some case series (Italy).^8^ But case reports from France and the UK have more acute presentation with shock with features of toxic shock syndrome(TSS).^2,9^ But these data are reported mostly from intensive care unit admissions rather than general pediatric population. The US data also showed atypical Kawasaki disease features in 40% of the patients.^4^ But this variability in clinical manifestations is difficult to distinguish on clinical ground. Out of 10 patients who had some form of CNS involvement, 5 of them had clinical seizures which add further to the wide clinical distribution of MIS-C. Chen TH in his review of 6 reports on MIS-C found that out of 187 children, 38% had neurological issues.^10^ The exact cause of this neurologic complication is not known, but it seems to be different from adult COVID 19 cases where the usual cause is cerebrovascular thromboembolism. Post infectious immune response might be responsible for neurologic manifestation.^10^Only 9 (42.8%) cases had respiratory distress at admission. One child had features of acute respiratory distress syndrome (ARDS) and rest 8 had features of either shock or pulmonary edema. Godfred-Cato S et al. in an USA cohort, had reported significant respiratory involvement (63%) in MIS-C. Those patients had higher incidence of RT-PCR positivity and higher mortality rate.^11^ In our series, the child who developed ARDS was found to be RTPCR positive and she expired despite all supportive care.

Out of 21 cases, 10 (48%) children presented with shock and required volume resuscitation and inotropic support. Echocardiography was done in 17 cases; myocardial dysfunction was found in 8 (38.1%), while we were not able to detect coronary dilatation in any patient. All the cases responded to the supportive treatment. Ramcharan T et al. in his cohort of 15 cases of MIS-C, who were referred for cardiac evaluation found cardiac dysfunction in 63% cases, while coronary abnormalities were detected in 93% cases.^12^ In a recent study from Mumbai, coronary affection was found in 23.8% cases.^13^

In the present study mean total leucocyte count is 13,800/cmm with a mean neutrophil 72%,lymphocyte 22% which is comparable to the Meta-analysis done by Lagunas-Range et al who could find an association between the high white cell count, low lymphocyte count, low platelet count, elevated CRP, and severity.^14^ In three patients, thrombocytopenia (less than 1.5 Lakh/cmm) was found but none had very low platelet count, which could be a point to distinguish this syndrome from severe sepsis or dengue. The inflammatory markers like CRP, ESR and procalcitonin were elevated in our enrolled patients with a mean CRP of 52.2 mg/dL. There is increasing evidence in adult studies, that raised levels of CRP are associated with severity and mortality in COVID patients.^15^Majority of our patients have shown positive IgG to SARS-CoV-2 which points towards an association with past COVID infections in these patients. In 2 cases where both RTPCR and antibodies were negative, the epidemiological connection with past COVID-19 infection could not be ignored, as their parents were RTPCT positive 4 weeks back. The mean D-dimer value was very high (3.832 mcg/ml) which is comparable to the elevated D-dimer levels in both pediatric and adult patients with COVID-19 ^16^. Evidence from adult studies suggests that elevated D-dimer levels are associated with poor outcome but therapeutic implications of the high D-dimer are yet to be understood fully.^17^ A recent pediatric guideline suggests thromboprophylaxis in children with COVID-19 with risk factors for thrombosis such as presence of central venous catheter, decreased mobility, and past or family history of thromboembolism.^18,19^American College of Rheumatology has recommended use of therapeutic anticoagulation for cases with ejection fraction less than 35%, documented thrombosis and coronary artery z score of more than 10.^20^ It is still not clear whether MIS-C patients have similar risk of thrombosis like acute COVID patients. However, as these patients have high D-dimer along with multi organ dysfunction requiring critical care treatment, it is reasonable to start low-molecular weight heparin (LMWH) for MIS-C.^21^We used LMWH in our patients as per the above guidelines. Myocardial dysfunction was found in 38.1%, while none had coronary dilatation in their first ECHO in our study.

Among 21 patients 9 received IVIG (1-2g/kg) but a higher percentage of patients received steroids. The decision of IVIG or steroid or both was taken based on clinical presentation, severity at presentation and status of cardiac involvement. Because of poor socioeconomic status, few patients were treated with steroid in place of IVIG, however they all improved. Due to overlap of clinical features, many clinical trials have proposed IVIG as first choice in treatment of MIS-C.^20^Whittaker et al reported 100% use for KD-like presentations, 72% for those with shock, and 61% for those with fever and inflammation alone.^3^ With such variation in management of MIS-C patients, there is suboptimal evidence to assess the superiority of various treatments. Steroid alone was used in 13 cases to a good effect. In our study 47.6% patients required inotropic support despite adequate fluid resuscitation. Early recognition of shock, judicious fluid resuscitation, appropriate initiation of inotropes and vasopressors are seemingly key factors for favourable outcome. Non-invasive ventilation was required in 2 children for respiratory distress with shock. Similarly, invasive ventilation was required for 2 children, one for a child with RTPCR positive with respiratory involvement, and the other one, a case of nephrotic syndrome with multi organ dysfunction. Both the children requiring invasive ventilation expired. In one of the largest cohort from US of 570 cases of MIS-C, 38.1% needed some form of respiratory support while in another case series from Mumbai, 39.1% required invasive respiratory support.^11,22^ Mortality (9%) of our study is higher to 1.8– 3% from western literature. None of our patient had coronary abnormality while 2 had mild cardiac dysfunction at discharge comparable to other studies.

## CONCLUSION

Multisystem inflammatory syndrome in children is a serious and life threatening disease in children which mandates multi-speciality involvement as cardiac and neurologic complications are significantly high. Diagnosis and management of MIS-C is a challenge as the guidelines are evolving and new data are coming up in literature. Pediatricians should familiarise themselves with the presentation of this new disease and start aggressive multispeciality management. Immunothearapy is the cornerstone in management; however, selection of the exact drug may need further studies.

## Data Availability

All data available

## Notes

### Competing Interest Statement

The authors have declared no competing interest.

### Funding Statement

no funding

### Author Declarations

Approved by IEC of KIMS, Bhubaneswar

